# Whole Exome Sequencing of Suspected Monogenic Cerebral Small Vessel Disease Patients reveals Novel Gene Associations

**DOI:** 10.64898/2026.02.08.26345868

**Authors:** Solomon. K. Guyler, Mohammed. M. Alfayyadh, Neven Maksemous, Rodney. A. Lea, Robert. A. Smith, Heidi. G. Sutherland, Lyn. R. Griffiths

## Abstract

Cerebral small vessel diseases (CSVDs) are a group of disorders affecting the small arteries, veins, and capillaries supplying the white matter and deep grey matter structures. They are the most common form of cerebrovascular disease, accounting for almost half of vascular dementia cases worldwide and approximately 20% of stroke incidence. Whilst genetic testing is a routine diagnostic tool for monogenic CSVDs, less than 20% of patients have a causal variant in known CSVD genes.

We performed whole exome sequencing on 117 patients suspected of monogenic CSVD that had previously tested negative for pathogenic variants in seven well-characterised CSVD genes (*NOTCH3, HTRA1, COL4A1, COL4A2, TREX1, GLA,* and *FOXC1).* Targeted analysis was conducted on known and associated CSVD genes as well as candidate genes causative of conditions with symptomatic overlap to CSVD. Burden analysis focusing on rare, functional variants was used to identify novel associations when compared against a cohort of 1035 non-neurological control samples.

We identified 18 candidate disease-causing variants across nine CSVD-associated genes and a significant burden of rare and rare, likely disease-causing heterozygous variants in *ABCC6*. Two genes identified from stroke and neurodegenerative disease gene panels, *MYH11* (adjusted *P*=1×10^-2^) and *NOTCH1* (adjusted *P*=1×10^-2^), also had a significant burden of candidate disease-causing variants. Additionally, we identified novel associations for seven genes (*COL7A1, HMCN1, LAMA1, MMP9, TENM4, TNC, TTN*) with monogenic CSVD. Our findings implicate several genes as potentially causal of monogenic CSVD, highlighting the need for more extensive genetic screening in suspected CSVD cases and also functional characterisation of novel implicated variants to determine their mechanistic role in CSVD pathogenesis.

## Introduction

Cerebral small vessel disease (CSVD) is a chronic, progressive condition that affects the small blood vessels of the brain. It is the most common form of cerebrovascular disease and includes a group of disorders that collectively represent the most prevalent monogenic cause of stroke, typically presenting with deep lacunar infarcts and widespread white matter changes (Chojdak-Łukasiewicz et al., 2021). The most common monogenic forms of CSVD are cerebral autosomal dominant arteriopathy with subcortical infarcts and leukoencephalopathy (CADASIL), caused by pathogenic variants in *NOTCH3*; CADASIL2, caused by heterozygous variants in *HTRA1*; Gould syndrome resulting from variants in *COL4A1/2*; and Fabry disease, caused by variants in *GLA* (Meschia et al., 2023). Genetic testing of CSVD genes is routine when a monogenic small vessel disease is suspected, however such screening only detects causal mutations in ∼20% of patients, suggesting that there are as yet unidentified genes contributing to monogenic CSVD (Dunn et al., 2020; Gorukmez et al., 2023).

Whole exome sequencing (WES) is a well characterised approach in the detection of novel associations between genes and Mendelian diseases and has been used to previously identify novel CSVD genes (Aloui et al., 2021; Rademakers et al., 2011). The Genomics Research Centre (GRC) has been conducting diagnostic genetic testing for CADASIL, the most common monogenic CVSD, since 1997. However, only 12-15% of suspected patients are identified with a causal variant in *NOTCH3* (Dunn et al., 2020; Guyler et al., 2025). The expansion of diagnostic testing to include several other monogenic CSVD genes (*HTRA1, COL4A1, COL4A2, GLA, TREX1, FOXC1*) resulted in an additional ∼14% improvement in the detection of likely damaging variants, yielding an overall diagnostic rate of 26%. Despite this, the majority of suspected patients remain without an identified causal mutation (Dunn et al., 2025). In this study, we performed WES on a cohort of 117 patients clinically suspected of monogenic CSVD. Novel associations were investigated using several bioinformatic methods and gene-based burden testing of candidate genes (Figure 1).

**Figure 1:**
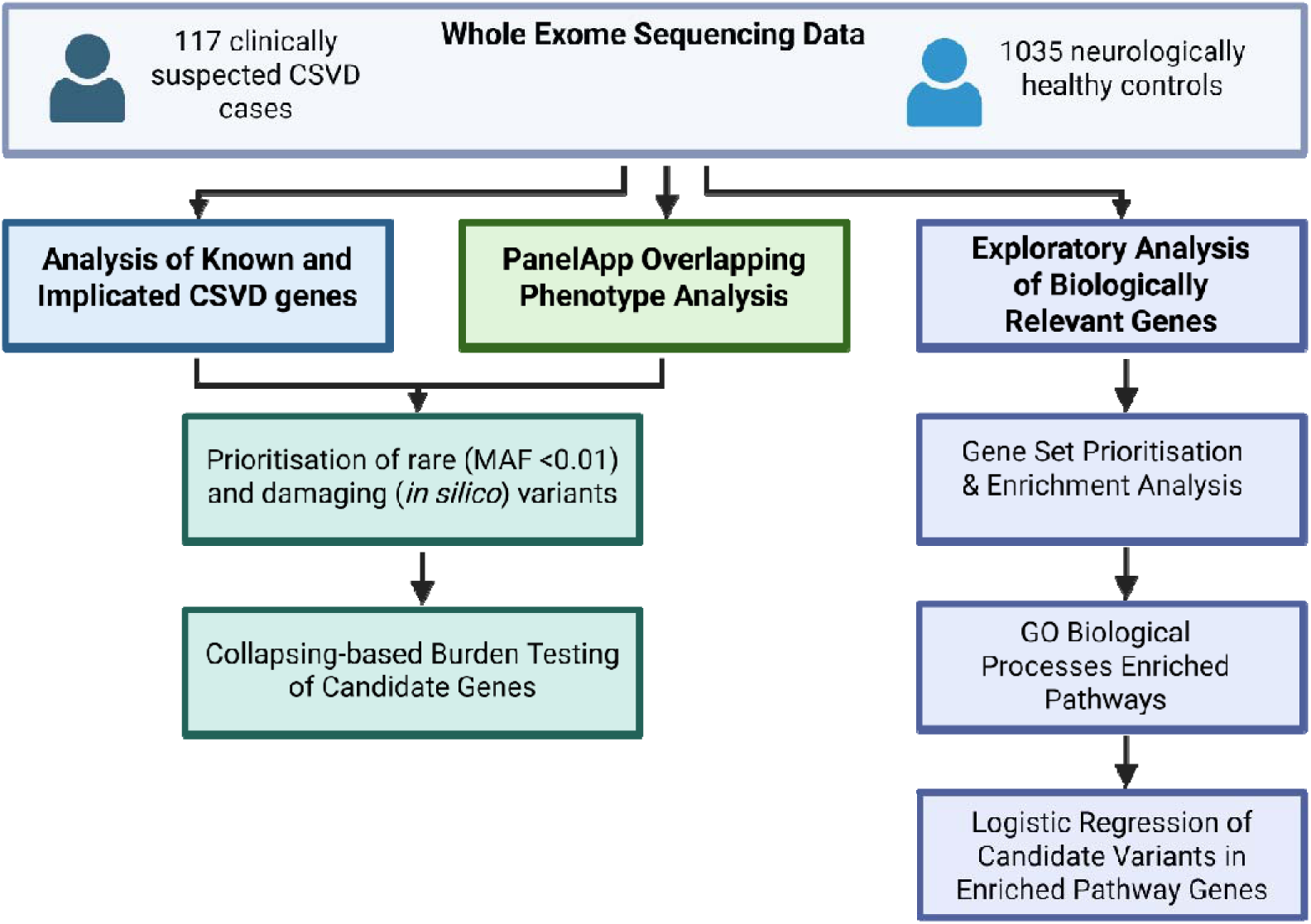
Study workflow. Incorporating WES data from our patient cohort and UKBB neurologically healthy controls, we first performed a targeted investigation of causal CSVD genes and genes associated with CSVD or cerebrovascular pathology. We then explored genes causative of alternate disorders with phenotypic overlap to monogenic CSVDs. Finally, we performed an exploratory analysis using gene set prioritisation and enrichment analyses to identify novel genetic associations with CSVD.

## Methods

### Patient Cohort

The study cohort included 117 patients referred to the Genomics Research Centre (Queensland University of Technology, Brisbane, Australia) for molecular genetic testing for monogenic CSVD according to National Association for Testing Authority, Australasia (NATA) standards. Participants were selected based upon the following inclusion criteria: (1) Clinical suspicion of CADASIL or other monogenic CSVD with clinical signs and symptoms consistent with CSVD, including recurrent stroke, evidence of vascular leukoencephalopathy or dementia, cognitive decline, migraine, or gait disturbance; and (2) negative for pathogenic variants in our targeted CSVD gene panel, which includes *NOTCH3, HTRA1, COL4A1, COL4A2, TREX1, FOXC1,* and *GLA*.

### Whole Exome Sequencing

DNA was extracted from peripheral blood using the QIAGEN QIACUBE™ according to the manufacturer’s instructions and quantified using the Nanodrop 8000 Spectrophotometer (Thermo Fisher Scientific). WES was performed using the Ion AmpliSeq Exome RDY-kits for library preparation according to the manufacturer’s instructions (MAN0010084). Library quantification was performed using an Invitrogen Qubit 3 Fluorometer (Thermo Fisher Scientific) for equimolar pooling prior to sequencing. Template preparation, enrichment and chip loading were performed using the Ion 550™ Kit-Chef (Cat. Number A34541) and 550 Chips on the Ion Chef. Sequencing was performed using the Ion S5 + platform with sequences aligned to human genome 38 (Hg38) and variant calling performed using PathVAR, a customisable NGS pipeline for whole exome/genome sequencing analysis and annotation (Alfayyadh et al., 2025). Coverage and mapping parameters showed an average coverage of 118X with a mean number of reads of 39,275,917. An average of 96% of reads were on target and the average >20X coverage was 89%.

### Control Cohort Generation

The control cohort used in this study was derived from the UK Biobank whole genome sequencing (WGS) resource, which includes genetic, biochemical, and imaging data for over 500,000 participants (Bycroft et al., 2018). From this cohort, we obtained data for a subset of 1,035 aged individuals (mean age = 53; SD = 7.3; 518 males, 517 females) without any history of neurological or cerebrovascular disease, serving as neurologically healthy controls. To equate the control dataset to the CSVD cohort, whole exome data for these individuals was extracted from WGS base alignment mapping (BAM) files using the view function in the BCFTools package (Danecek et al., 2021).

### Estimation of Relatedness and Linkage

Underlying population structure and relatedness were assessed through principal component analysis (PCA) of genotypes shared between our case and control cohort and the 1,000 Genome Project Phase 3 data (Zheng-Bradley et al., 2017). 12122 single nucleotide polymorphisms (SNPs) were selected after removing all SNPs with a minor allele frequency (MAF) of ≤0.05. SNPs in linkage disequilibrium were pruned using PLINK1.9 software with a window size of 50bp, a window shift of 5bp, and a variant inflation factor of 0.1 (Purcell et al., 2007). Filtered samples were then used to perform a PCA using PLINK1.9. Related individuals were identified using pairwise identity-by-descent (IBD) distances, and kinship values were generated and extracted with samples with an IBD >0.1 excluded from downstream analysis. Of the 117 participants in the case cohort, 94.8% were assigned to European ancestry (n=111), 2.56% to South Asian (n=3), and 2.56% to East Asian (n=3). All control samples (n=1035) from the UKBB were assigned European ancestry and included in downstream burden analysis.

### Variant Filtering

Variants were filtered based on coverage and allele ratio with a minimum coverage of 20x and an allele ratio of 35% to 65% used for heterozygous variants and >85% used for homozygous variants. Variants predicted to alter protein function, including frameshift, non-synonymous, stop-gain, and stop-loss variants, were retained. Population frequency filtering was applied using gnomAD v4.1.0, with variants observed at a MAF ≤0.01 in non-Finnish Europeans retained for downstream analysis. Functional impact was assessed using *in-silico* prediction tools, including Polymorphism Phenotyping v2 (PolyPhen-2), Sorting Intolerant From Tolerant (SIFT4G), REVEL, and Combined Annotation Dependent Depletion (CADD v1.7) (Adzhubei et al., 2013; Ioannidis et al., 2016; Rentzsch et al., 2019; Sim et al., 2012). Variants classified as deleterious by at least two tools using score guidelines in accordance with American College of Medical Genetics (ACMG) guidelines were retained as candidate variants for burden testing (Pejaver et al., 2022). Variants were further assessed using ClinVar with variants classified as benign or likely benign excluded as not disease-causing.

### Burden Testing

Burden testing was performed using a gene-based collapsing framework. For each gene in the lists outlined below, individuals carrying at least one candidate variant were compared between cases and controls. Statistical testing was performed for known and associated CSVD genes and PanelApp candidate genes using Fisher’s exact test, implemented in R, while logistic regression using the glmnet package in R was used for the exploratory analysis, with age and sex included as covariates in the regression model (Friedman et al., 2010). A nominal significance threshold of *P* < 0.05 was applied, and correction for multiple testing was performed using the Benjamin–Hochberg false discovery rate (FDR) (Noble, 2009). Genes with an adjusted *P* < 0.05 were considered statistically significant.

### Known and Implicated CSVD Gene Analysis

Analysis was performed on known and associated CSVD genes and genes implicated in cerebrovascular pathology (Fig. 2) (Aloui et al., 2021; Bordes et al., 2022; Ding et al., 2023; Miyatake et al., 2018; Nicholson et al., 2013). Candidate rare, likely disease-causing variants were identified following filtering and classification steps described above.

**Figure 2:**
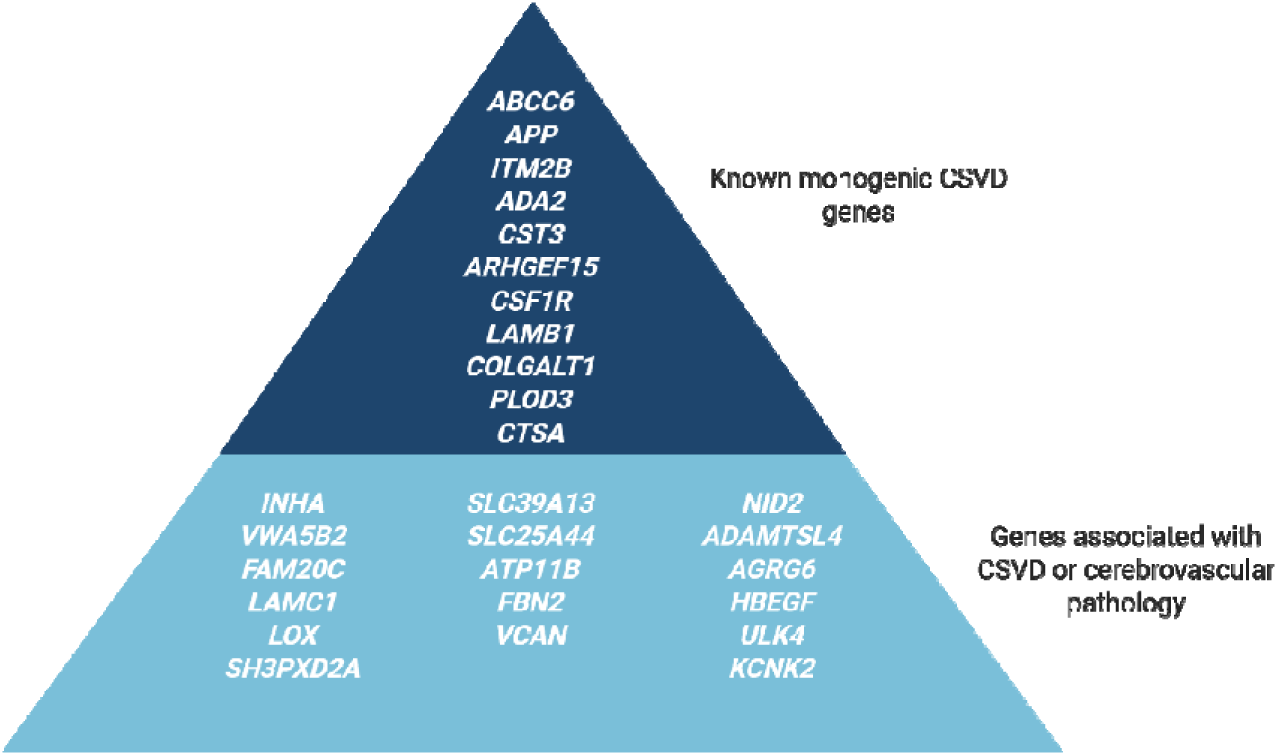
Gene list used for rare variant analysis. Genes were identified as either causal of a distinct monogenic CSVD or as associated through burden testing, GWAS, TWAS, or functional analysis identifying a role in driving cerebrovascular pathology.

### PanelApp Gene Set Analysis

Targeted analysis was conducted using gene panels with potential overlap to CSVD which were curated in PanelApp Australia, including Stroke (v1.16), Cerebral vascular malformations (v1.0), Leukodystrophy – adult-onset (v0.147), Neurodegenerative disease – adult-onset (v6.118), and Cerebral amyloid angiopathy (v1.1) (Table S1) (Stark et al., 2021).

### Gene Set Enrichment Analysis

A gene set was curated where genes containing at least one rare, likely disease-causing variant were collated. This gene set was prioritised using ToppGene Suite (default parameters), with known CSVD genes (*NOTCH3, HTRA1, COL4A1, COL4A2, GLA, TREX1, CTSA, FOXC1, ABCC6, COLGALT1, APP, CST3, ITM2B, ADA2*) used as the training set (Chen et al., 2009). The ranked list of genes was then subjected to gene set enrichment analysis (GSEA) using WebGestalt (default parameters), focusing on Gene Ontology Biological Processes to identify pathways relevant to CSVD pathogenesis (Elizarraras et al., 2024). Genes identified in significantly enriched pathways from GSEA were obtained, resulting in a list of candidate genes for association analysis using logistic regression (Table S2). Genes used within the training set for gene set prioritisation were excluded from this analysis due to their established role in CSVD. Significantly associated genes were retained for further evaluation.

### Gene Expression Analysis

Gene expression data was obtained from the Genotype-Tissue Expression (GTEx) portal and the CELLxGENE Discover corpus (Jones et al., 2022; Lonsdale et al., 2013). Expression was assessed across brain regions and vascular cell types. Aggregated expression across tissues was calculated, and genes were assessed by scaled expression and percentage tissue composition.

## Results

### Analysis of Known and Recently Implicated CSVD Genes

117 patients suspected of monogenic CSVD underwent WES to identify potential causes of disease. Twenty-eight genes previously identified as causal or associated with monogenic CSVD were assessed for causal variants in this cohort (Figure 2). 75 rare variants were identified in 19 genes, with some occurring in multiple individuals (Table S3). No variants were detected in *FAM20C*, *CST3*, *ITM2B, LOX, AGRG6, KCNK2, SLC39A13, SLC25A44,* or *CTSA*.

When restricting to variants predicted to be damaging by *in-silico* tools, 18 rare, likely disease-causing variants were identified in the CSVD cohort across *ABCC6, LAMB1, APP, COLGALT1 LAMC1, EFEMP1, FBN2, SH3PXD2A,* and *VCAN* (Table I). Five heterozygous variants were identified in *ABCC6*, including two pathogenic variants and three classified as variants of uncertain significance (VUS). One heterozygous VUS variant was identified in *COLGALT1*, a gene typically associated with autosomal recessive CSVD. All other variants identified were classified as VUS or had not previously been annotated within ClinVar.

**Table I:** Rare, Likely Disease-Causing Variants Identified in Known and Implicated CSVD Genes.

| Gene | Chromosome Position | Variant | Patient | Minor Allele Frequency | ClinVar Classification | CADD | Poly Phen | SIFT | REVEL |
| --- | --- | --- | --- | --- | --- | --- | --- | --- | --- |
| **ABCC6** | chr16:16163078 | c.3421C>T<br>p.Arg1141Ter | DGR545 | 0.001943 | Pathogenic | 42 | N/A | N/A | - |
| **ABCC6** | chr16:16202006 | c.1171A>G<br>p.Arg391Gly | DGR25 | 0.007767 | Pathogenic/VUS/Likely Benign | 22.9 | 1 | 0.002 | 0.847 |
| **ABCC6** | chr16:16182476 | c.2183C>G<br>p.Ala728Gly | DGR569 | 0.00001695 | VUS | 23.4 | 0.95 | 0.004 | 0.739 |
| **ABCC6** | chr16:16169823 | c.2818C>T<br>p.Arg940Cys | DGR271 | 8.68E-07 | VUS | 23.9 | 0.591 | 0 | 0.612 |
| **ABCC6** | chr16:16157692 | c.3853G>A<br>p.Val1285Met | DGR363 | 0.00003305 | VUS | 22 | 0.591 | 0.004 | 0.477 |
| **APP** | chr21:26000072 | c.976G>A<br>p.Gly326Ser | DGR349 | 0.00002288 | VUS | 27.7 | 0.979 | 0 | 0.591 |
| **APP** | Chr21:26000019 | c.1027A>C<br>p.Ser343Arg | DGR474 | 8.475E-07 | N/A | 25.1 | 0.741 | 0 | 0.334 |
| **LAMB1** | chr7:107975088 | c.1380C>A<br>p.Cys460Ter | DGR24 | 0.000001707 | N/A | 39 | N/A | N/A | - |
| **LAMB1** | chr7:107980808 | c.680T>C<br>p.Leu227Ser | DGR330 | 0.001144 | VUS/Likely Benign | 24.2 | 0.702 | 0.001 | 0.682 |
| **LAMB1** | chr7:107959319 | c.2620G>T<br>p.Asp874Tyr | DGR556 | N/A | N/A | 23.7 | 0.982 | 0.01 | 0.407 |
| **LAMC1** | chr1:183127313 | c.3032G>A<br>p.Arg1011His | DGR524 | 0.006805 | N/A | 25.6 | 0.987 | 0.006 | 0.592 |
| **LAMC1** | chr1:183118125 | c.1969C>A<br>p.Arg657Ser | DGR531 | 0.0001153 | N/A | 28.1 | 1 | 0 | 0.625 |
| **LAMC1** | chr1:183110594 | c.961C>T<br>p.Pro321Ser | DGR558 | 0.001071 | N/A | 25.7 | 0.975 | 0.018 | 0.374 |
| **COLGALT1** | chr19:17692146 | c.1762C>T<br>p.Arg588Cys | DGR75 | 0.0003288 | VUS | 27.2 | 1 | 0.011 | 0.731 |
| **EFEMP1** | chr2:55881684 | c.568T>C<br>p.Cys190Arg | DGR24 | 0.000005085 | VUS | 27.8 | 0.999 | 0 | 0.974 |
| **FBN2** | chr5:128263603 | c.8014A>G<br>p.Thr2672Ala | DGR538 | 8.457E-7 | VUS | 27.7 | 0.996 | 0 | 0.877 |
| **SH3PXD2A** | chr10:103735778 | c.260G>A<br>p.Arg87Gln | DGR342 | 0.000005932 | VUS | 26.6 | 0.96 | 0 | 0.293 |
| **VCAN** | chr5:83512306 | c.952G>T<br>p.Gly318Cys | DGR27 | 0.00002882 | VUS | 29.5 | 0.999 | 0 | 0.372 |

*The chromosomal assembly used in this analysis was human genome 38 (hg38). Minor allele frequencies were obtained from the GnomAD v4.1.0 non-Finnish European population. Transcript identifiers: LAMB1 (NM\_002291.3), APP (NM\_000484.4), ABCC6 (NM\_001171.6), LAMC1 (NM\_002293.3), COLGALT1 (NM\_024656.3), EFEMP1 (NM\_001039348.3), FBN2 (NM\_001999.4), SH3PXD2A (NM\_001394015.1), and VCAN (NM\_004385.5).*

Principal component analysis was performed for ancestry inference, and six cases were excluded due to non-European ancestry, leaving 111 CSVD cases for burden analysis. Burden testing of these genes using a non-neurological control cohort from the UKBB (n=1035) identified a significant association with the CSVD cohort for *ABCC6*, both when considering rare variants (adjusted *P* = 9.42 × 10⁻□) and when restricting to rare, likely damaging variants (adjusted *P* = 0.011) (Table II). A significant association was also observed for rare *FBN2* variants which were enriched in the control cohort (adjusted *P* = 0.041, OR = 0.30), indicating a lower frequency of *FBN2* variants among individuals with CSVD.

**Table II:**
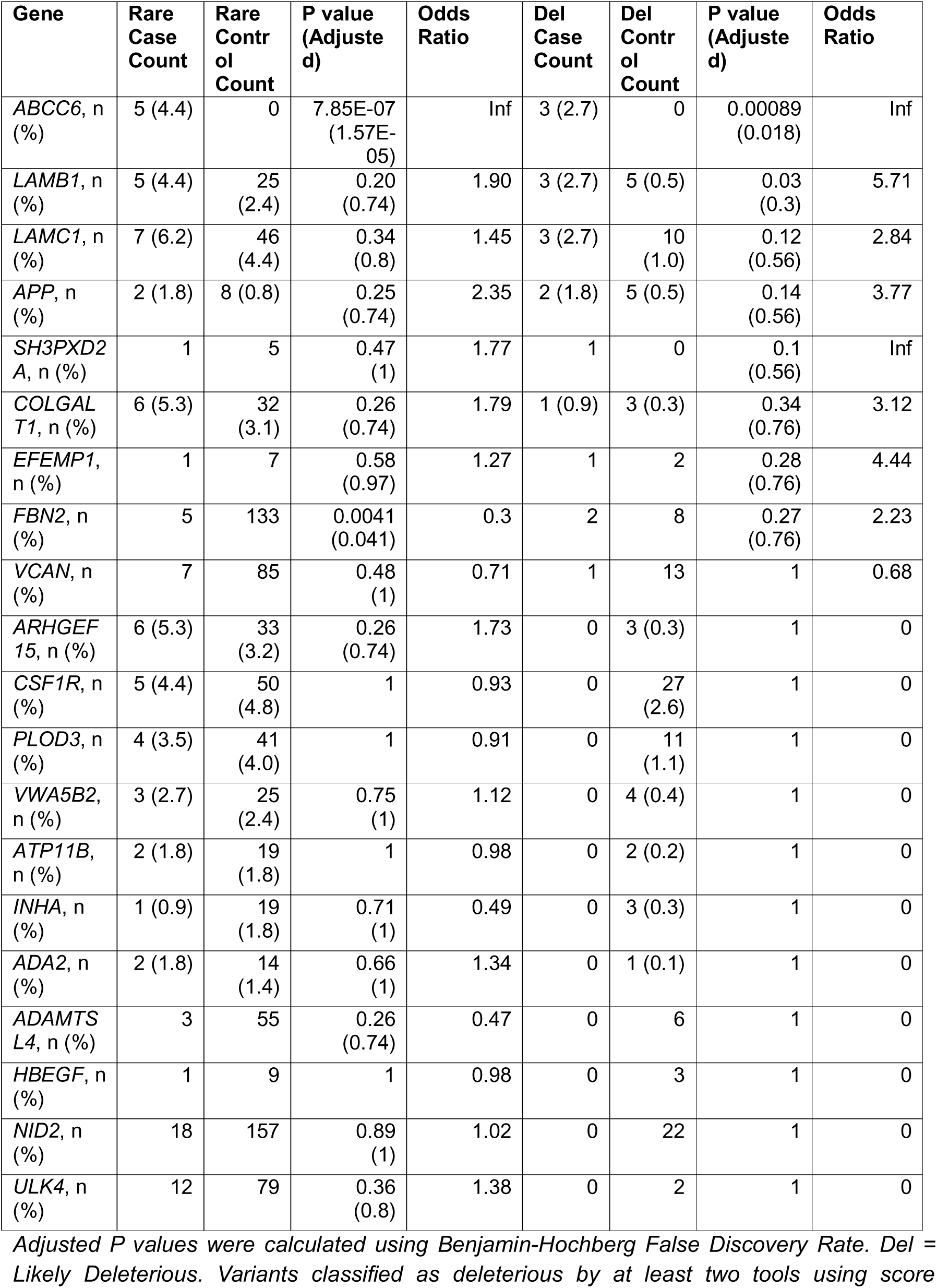
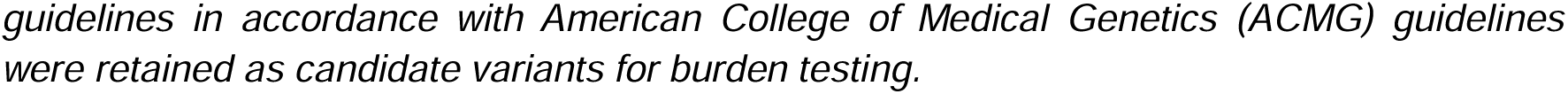
Fisher’s Exact Test Comparing the Burden of Rare Variants and Rare Likely Disease-causing Variants in Known and Implicated CSVD Genes.

| Gene | Rare Case Count | Rare Control Count | P value (Adjusted) | Odds Ratio | Del Case Count | Del Control Count | P value (Adjusted) | Odds Ratio |
| --- | --- | --- | --- | --- | --- | --- | --- | --- |
| ABCC6, n (%) | 5 (4.4) | 0 | 7.85E-07 (1.57E-05) | Inf | 3 (2.7) | 0 | 0.00089 (0.018) | Inf |
| LAMB1, n (%) | 5 (4.4) | 25 (2.4) | 0.20 (0.74) | 1.90 | 3 (2.7) | 5 (0.5) | 0.03 (0.3) | 5.71 |
| LAMC1, n (%) | 7 (6.2) | 46 (4.4) | 0.34 (0.8) | 1.45 | 3 (2.7) | 10 (1.0) | 0.12 (0.56) | 2.84 |
| APP, n (%) | 2 (1.8) | 8 (0.8) | 0.25 (0.74) | 2.35 | 2 (1.8) | 5 (0.5) | 0.14 (0.56) | 3.77 |
| SH3PXD2A, n (%) | 1 | 5 | 0.47 (1) | 1.77 | 1 | 0 | 0.1 (0.56) | Inf |
| COLGALT1, n (%) | 6 (5.3) | 32 (3.1) | 0.26 (0.74) | 1.79 | 1 (0.9) | 3 (0.3) | 0.34 (0.76) | 3.12 |
| EFEMP1, n (%) | 1 | 7 | 0.58 (0.97) | 1.27 | 1 | 2 | 0.28 (0.76) | 4.44 |
| FBN2, n (%) | 5 | 133 | 0.0041 (0.041) | 0.3 | 2 | 8 | 0.27 (0.76) | 2.23 |
| VCAN, n (%) | 7 | 85 | 0.48 (1) | 0.71 | 1 | 13 | 1 | 0.68 |
| ARHGEF15, n (%) | 6 (5.3) | 33 (3.2) | 0.26 (0.74) | 1.73 | 0 | 3 (0.3) | 1 | 0 |
| CSF1R, n (%) | 5 (4.4) | 50 (4.8) | 1 | 0.93 | 0 | 27 (2.6) | 1 | 0 |
| PLOD3, n (%) | 4 (3.5) | 41 (4.0) | 1 | 0.91 | 0 | 11 (1.1) | 1 | 0 |
| VWA5B2, n (%) | 3 (2.7) | 25 (2.4) | 0.75 (1) | 1.12 | 0 | 4 (0.4) | 1 | 0 |
| ATP11B, n (%) | 2 (1.8) | 19 (1.8) | 1 | 0.98 | 0 | 2 (0.2) | 1 | 0 |
| INHA, n (%) | 1 (0.9) | 19 (1.8) | 0.71 (1) | 0.49 | 0 | 3 (0.3) | 1 | 0 |
| ADA2, n (%) | 2 (1.8) | 14 (1.4) | 0.66 (1) | 1.34 | 0 | 1 (0.1) | 1 | 0 |
| ADAMTS L4, n (%) | 3 | 55 | 0.26 (0.74) | 0.47 | 0 | 6 | 1 | 0 |
| HBEGF, n (%) | 1 | 9 | 1 | 0.98 | 0 | 3 | 1 | 0 |
| NID2, n (%) | 18 | 157 | 0.89 (1) | 1.02 | 0 | 22 | 1 | 0 |
| ULK4, n (%) | 12 | 79 | 0.36 (0.8) | 1.38 | 0 | 2 | 1 | 0 |

Adjusted P values were calculated using Benjamin-Hochberg False Discovery Rate. Del = Likely Deleterious. Variants classified as deleterious by at least two tools using score
*guidelines in accordance with American College of Medical Genetics (ACMG) guidelines were retained as candidate variants for burden testing.*

### Investigation of Candidate Genes from PanelApp Australia Gene Panels

Genes included in PanelApp Australia gene panels, a publicly available diagnostic gene panel resource, which were related to neurodegenerative disease, stroke, cerebrovascular malformation, or leukodystrophy were analysed. Variants in genes previously implicated in CSVD and examined in the initial analysis of CSVD-associated genes were excluded from this analysis. Rare, likely disease-causing variants were identified in 35 of 111 individuals (31.5%), all of which were heterozygous. In total, 34 unique variants were observed across 24 genes, including 30 missense variants and four nonsense variants introducing premature stop codons (Table S4). Four patients were identified with pathogenic variants, including three patients with a pathogenic variant in *SQSTM1*, causative of frontotemporal dementia with amyotrophic lateral sclerosis, and one patient carrying a nonsense variant in *ANGPTL6* which has been shown to cause familial intracranial aneurysm (Table III).

**Table III:** Disease-causing variants identified in genes associated with alternate disorders.

| Patient | Chromosome Position | Gene | Variant | Minor Allele Frequency | ClinVar Classification | CAD D | Poly Phen | SIFT | REVEL |
| --- | --- | --- | --- | --- | --- | --- | --- | --- | --- |
| **DGR350**<br>**DGR527** | chr5:179836445 | *SQSTM1* | c.1175C>T<br>p.Pro392Leu | 0.001674 | Pathogenic/Likely pathogenic/VUS | 24.7 | 0.3 | 0.07 | 0.818 |
| **DGR565** | chr5:179823038 | *SQSTM1* | c.286C>T<br>p.Arg96Ter | 0.00001356 | Pathogenic | 44 | N/A | N/A | - |
| **DGR524** | chr19:10093793 | *ANGPTL6* | c.851C>G<br>p.Ser284Ter | 0.0007635 | N/A | 36 | N/A | N/A | - |

*The chromosomal assembly used in this analysis was human genome 38 (hg38). Minor allele frequencies were obtained from the GnomAD v4.1.0 non-Finnish European population. Transcript identifiers: SQSTM1 (NM\_003900.5), ANGPTL6 (NM\_031917.3). VUS: variant of uncertain significance.*

Burden testing was performed on all genes from the selected PanelApp gene panels containing at least one rare, likely disease-causing variant. Significant associations with the CSVD cohort were identified for two genes, *MYH11* and *NOTCH1,* compared with controls (Figure 3). Despite there being nine times as many controls as cases, no candidate variants in these genes were detected in the controls compared with three variants in each gene in the case cohort (Table IV). Notably, likely disease-causing variants in each of these genes were also identified in patients who had previously been excluded from burden analysis due to non-European ancestry (Table S5). Comprehensive details of burden testing results are available in Table S6.

**Figure 3:**
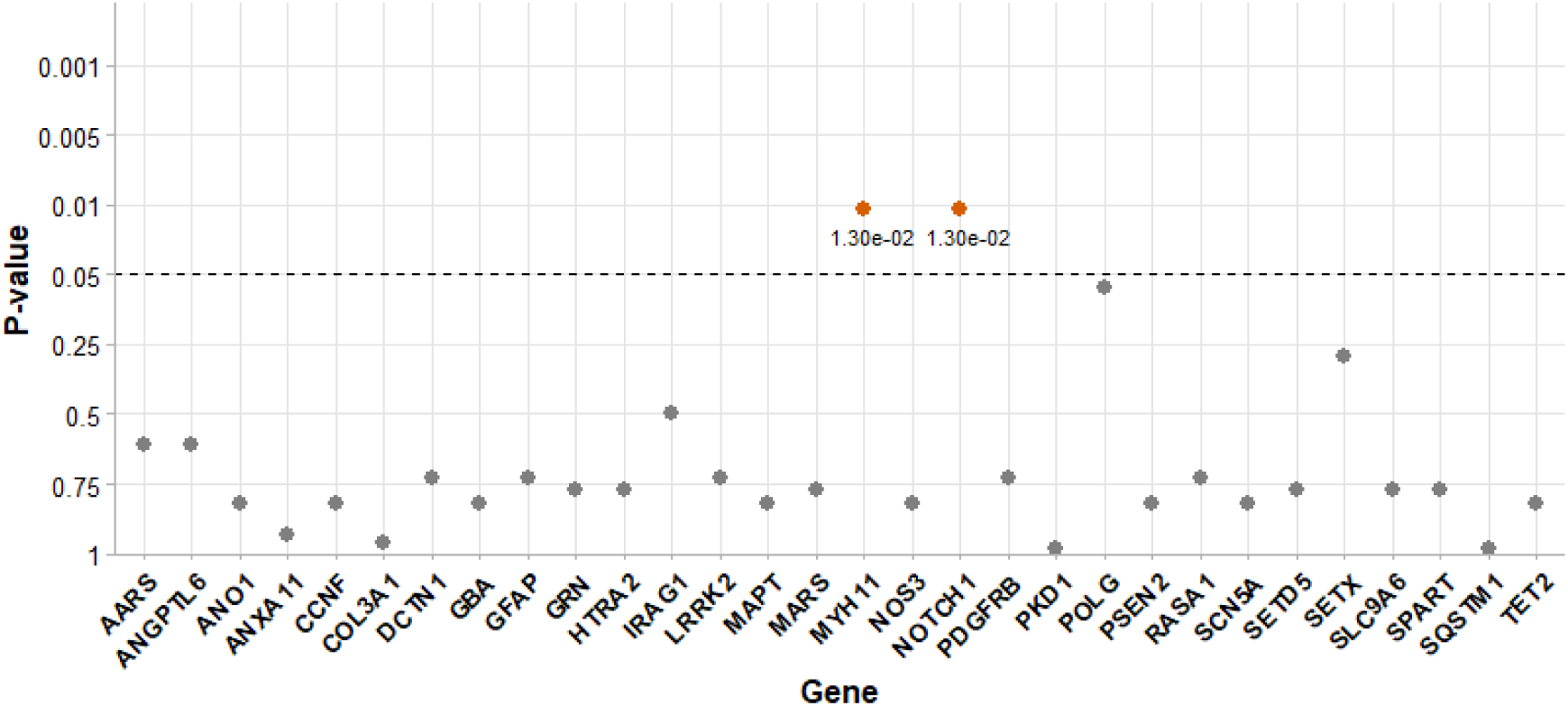
Gene-based Burden Testing Results from Genes Included in Candidate PanelApp Australia Gene Panels. P values shown have been adjusted using the Benjamin-Hochberg False Discovery Rate Method. The dotted line indicates the statistical significance threshold of P = 0.05.

**Table IV:** Disease-causing variants identified in significant genes from PanelApp gene panels with phenotypic overlap to CSVD.

| Patient | Chromosome Position | Gene | Variant | Minor Allele Frequency | ClinVar Classification | CAD D | Poly Phen | SIFT | REVEL |
| --- | --- | --- | --- | --- | --- | --- | --- | --- | --- |
| **DGR22** | chr16:15718337 | *MYH11* | c.5294G>A<br>p.Arg1765Gln | 0.0002449 | Likely Pathogenic/VUS/Likely Benign | 29.9 | 0.982 | 0.033 | 0.864 |
| **DGR540** | chr16:15720973 | *MYH11* | c.4678G>T<br>p.Asp1560Tyr | - | - | 31 | 1 | 0 | 0.833 |
| **DGR278** | chr16:15778801 | *MYH11* | c.790G>A<br>p.Val264Met | 0.00015 | VUS | 22.8 | 0.857 | 0.003 | 0.717 |
| **DGR538** | chr9:136508893 | *NOTCH1* | c.3148G>A<br>p.Gly1050Ser | 0.000006105 | VUS | 26.9 | 1 | 0 | 0.858 |
| **DGR32** | chr9:136506782 | *NOTCH1* | c.3835C>T<br>p.Arg1279Cys | 0.0009145 | VUS/Likely Benign/Benign | 29.7 | 0.841 | 0.063 | 0.436 |
| **DGR307** | chr9:136510659 | *NOTCH1* | c.2734C>T<br>p.Arg912Trp | 0.003148 | VUS/Likely Benign/Benign | 25.4 | 0.85 | 0.017 | 0.402 |

*The chromosomal assembly used in this analysis was human genome 38 (hg38). Minor allele frequencies were obtained from the GnomAD v4.1.0 non-Finnish European population. Transcript identifiers: MYH11 (ENST00000396324.7), NOTCH1 (NM\_017617.5). VUS: variant of uncertain significance.*

### Identification of Novel Monogenic CSVD Genes

Finally, we performed analyses using all SNVs across 117 WES samples to identify novel genetic associations with CSVD. In the WES CSVD dataset, we identified 5,527 rare, damaging variants across 3,419 unique genes. Genes carrying at least one rare, damaging variant were then ranked by biological and functional similarity to established CSVD genes. Gene set enrichment analysis (GSEA) focusing on GO: Biological Processes identified ten significantly enriched pathways, with endoderm development and morphogenesis of a branching structure identified as the most significant (Table S7). A total of 221 unique genes were identified from GSEA enriched pathways and retained for burden analyses, excluding genes previously established as causal or associated with CSVD. Seven genes (*TTN, TENM4, COL7A1, MMP9, HMCN1, LAMA1,* and *TNC*) showed significant associations with CSVD (Table V).

**Table V:** Significant Genes Identified from Burden Testing of Rare, Deleterious Variants in Genes Identified in GO: Biological Process-Enriched Pathways.

| Gene | Variant Carriers in cases (% of cohort) | Variant Carriers in controls (% of cohort) | P Value (Adjusted) | Odds Ratio |
| --- | --- | --- | --- | --- |
| **COL7A1, n (%)** | 6 (5.3) | 4 (0.4) | 0.000567<br>(0.03377) | 10.646 |
| **HMCN1, n (%)** | 4 (3.5) | 4 (0.4) | 0.000838<br>(0.03663) | 11.424 |
| **LAMA1, n (%)** | 8 (7.1) | 14 (1.4) | 0.000999<br>(0.03663) | 4.863 |
| **MMP9, n (%)** | 4 (3.5) | 3 (0.3) | 0.000614<br>(0.03377) | 14.743 |
| **TENM4, n (%)** | 6 (5.3) | 9 (0.9) | 0.000244<br>(0.02684) | 7.478 |
| **TNC, n (%)** | 4 (3.5) | 3 (0.3) | 0.00157<br>(0.04934) | 11.804 |
| **TTN, n (%)** | 14 (12.4) | 31 (3.0) | 0.0000277<br>(0.006094) | 4.354 |

*Burden analysis was performed using logistic regression with age and sex used as covariates. A nominal significance threshold of $P = 0.05$ after multiple testing correction was set.*

Gene expression analysis of genes identified as associated with CSVD was performed using GTEx and CELLxGENE. GTEx showed *MYH11, TNC, NOTCH1, COL7A1,* and *HMCN1* as highly expressed in arterial tissues, while *NOTCH1* showed the highest expression across brain tissues amongst the genes tested (Figure 4). Single-cell RNA expression data from CELLxGENE revealed *TENM4* and *TNC* as having the highest aggregate expression in brain tissues, while *MYH11, HMCN1,* and *TNC* showed the highest expression in vascular cell types (Figure S1). Detailed information on all genes included in burden testing is provided in Table S8 and individual variants identified in significant genes are provided in Table S9.

**Figure 4:**
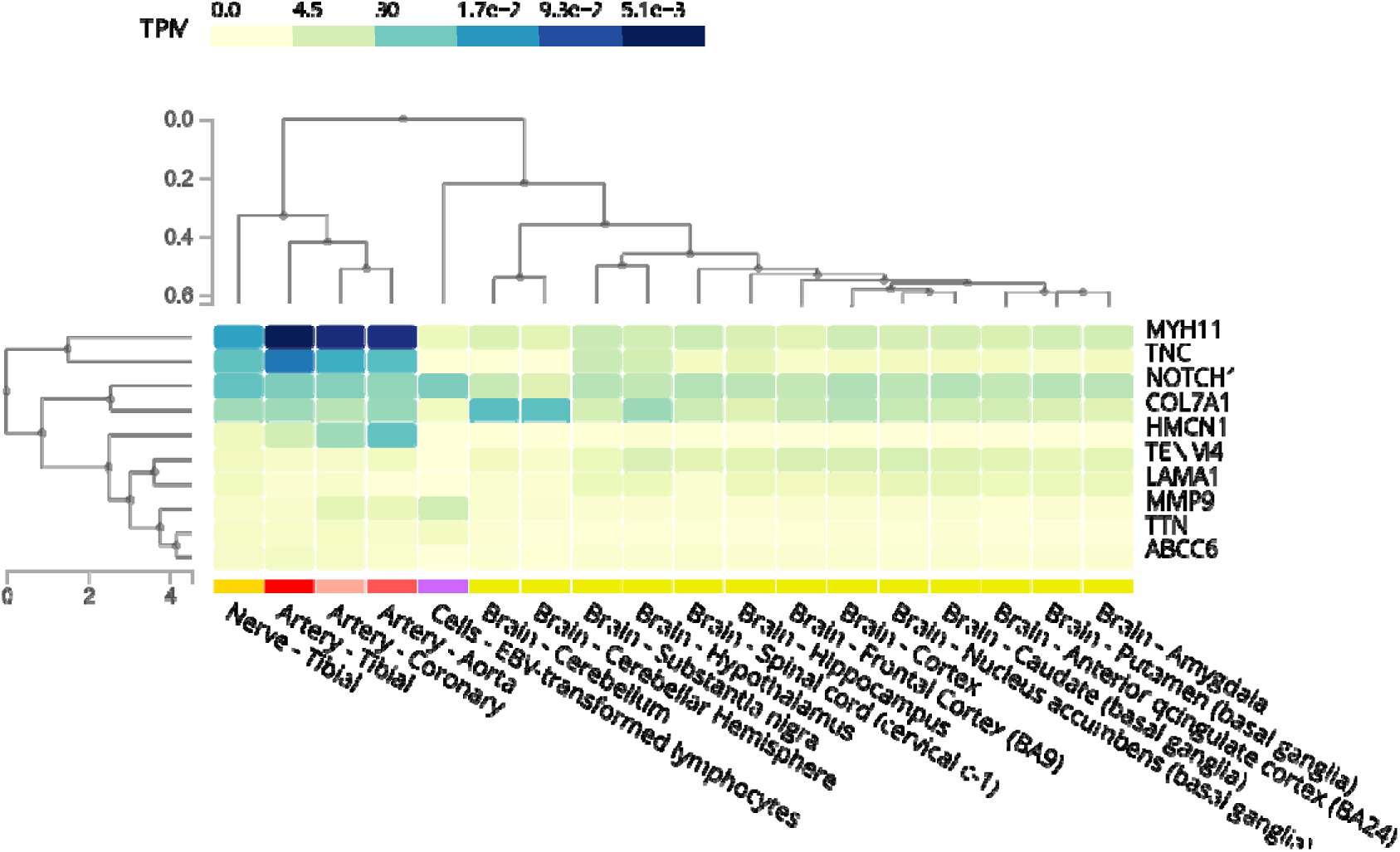
Gene Expression Map of Vasculature and Brain Tissues for Significant Genes Identified from PanelApp Gene Panels and Gene Set Enrichment Analysis.

## Discussion

In a cohort of 117 cases and 1,035 controls, we investigated damaging variants and gene-based associations with monogenic CSVD. A total of 15.4% of cases carried a candidate disease-causing variant in a known or recently associated CSVD gene. Additionally, we identified a significant burden of rare, deleterious variants in ten genes, including *ABCC6, NOTCH1, MYH11, TTN, TENM4, TNC, MMP9, HMCN1, LAMA1,* and *COL7A1*.

Five heterozygous variants were identified in *ABCC6,* causal of pseudoxanthoma elasticum (PXE), an autosomal recessive connective tissue disorder characterised by fragile vasculature, skin laxity, and retinal haemorrhage (Germain, 2017; Manini & Pantoni, 2021). Two of these variants have previously been identified as pathogenic in a recessive inheritance pattern, however monoallelic variants have been associated with a milder form of disease known as PXE forme fruste. The vascular wall degeneration and abnormal extra cellular matrix (ECM) remodelling described in PXE overlaps with CSVD pathogenesis and symptoms, including arterial stiffening, calcification of small and medium-sized vessels leading to transient ischaemic attack or stroke, white matter lesions, and cognitive decline (Kim et al., 2023; Legrand et al., 2017; Sunmonu, 2021). Whilst a fully penetrant PXE is considered an autosomal recessive disorder, heterozygous variants have been associated with prominent PXE-related cardiovascular phenotypes (Jin et al., 2021; Martin et al., 2008). Additionally, heterozygous pathogenic variants in *ABCC6* are associated with an increased risk of ischaemic stroke (De Vilder et al., 2018). A significant burden of both rare and predicted deleterious heterozygous variants was identified in *ABCC6*, highlighting its role as a potential cause of monogenic CSVD in this cohort.

Analysis of genes causal of disorders sharing CSVD symptomology identified a significant burden of rare, disease-causing variants in *NOTCH1* and *MYH11*. Three variants were detected in *NOTCH1*, a transmembrane receptor essential for vascular mural cell development (Zhou et al., 2022). While traditionally associated with congenital disorders such as Adams-Oliver disease through a loss-of-function mechanism, gain-of-function variants have been identified to cause a chronic CNS disorder with calcifying microangiopathy and leukoencephalopathy (Cho et al., 2022; Della Vecchia et al., 2024; Helman et al., 2022; Stanley et al., 2024). All reported pathogenic variants for this CNS disorder localize to the negative regulatory region (NRR), whereas variants identified in this study were in the epidermal growth factor-like repeat (EGFr) domains, highlighting the need for functional studies to determine potential converging mechanisms of pathogenesis.

*MYH11* encodes a myosin heavy chain subunit and is associated with non-syndromic thoracic aortic aneurysm and dissection (TAAD) (Pannu et al., 2007). Three candidate variants were identified in our case cohort, including one variant (p.Arg1765Gln) identified in DGR22 which has previously been classified as likely pathogenic for TAAD. Several cases of cerebral vascular pathologies caused by variants in *MYH11* have been identified, particularly as a cause of recurrent stroke in paediatric and young adult populations (Petrea et al., 2025; Raghuram et al., 2023). The established role of *MYH11* variants in vascular smooth muscle dysfunction and its association with vascular arteriopathy suggest that cerebral manifestations may also fall within the phenotypic spectrum of *MYH11*-related disorders (Atash et al., 2025).

Two pathogenic variants were also identified in three patients in *SQSTM1,* which is causative of frontotemporal dementia and/or amyotrophic lateral sclerosis (FTD-ALS) (Rubino et al., 2012). FTD-ALS shares clinical signs and symptoms with CSVD including gait disturbance, cognitive decline, and white matter changes, and several studies have identified clinical signs of ALS in patients with genetically diagnosed CVSD (Kim et al., 2012; Praline et al., 2010). An additional pathogenic nonsense variant was identified in *ANGPTL6* which is causative of familial intracranial aneurysm (fIA) (Bourcier et al., 2018). This variant has previously been identified in one patient with fIA and has been shown to result in complete loss of secretion of ANGPTL6 in cells (Hendricks et al., 2017; Hostettler et al., 2021).

Our burden analysis further identified significant associations between CSVD and seven additional genes: *TTN, TENM4, COL7A1, MMP9, HMCN1, LAMA1*, and *TNC. LAMA1* implicates a role for the ECM laminins in CSVD, particularly due to their role in CNS blood vessel integrity (Halder et al., 2023). *LAMA1* encodes the laminin α1 subunit and is critical to basement membrane architecture. Biallelic variants in *LAMA1* are associated with Poretti–Boltshauser syndrome, which causes cerebellar dysplasia, ocular abnormalities, and cognitive dysfunction (Powell et al., 2021). Gene knockdown of *LAMA1* in zebrafish models has shown hyaloid and trunk vasculature defects similar to the vascular defects identified in dual zebrafish *FOXC1a* and *FOXCb* knockout models (Edwards et al., 2010). These paralogs of the human *FOXC1*, a gene causative of autosomal dominant CSVD with prominent ocular features, highlight a potential link between *LAMA1* dysfunction and cerebrovascular disease (French et al., 2014).

Three variants were identified in *LAMB1* in our cohort which encodes the laminin β1 subunit. Protein-truncating variants in *LAMB1* that escape nonsense-mediated decay (NMD) have been reported to cause a monogenic CSVD characterised by hippocampal memory deficits and leukoencephalopathy (Aloui et al., 2021). Two missense substitutions (p.Leu227Ser and p.Asp874Tyr) and one nonsense variant (p.Cys460Ter) were identified in our cohort. Previous studies have identified cysteine-altering mutations as causative of leukoencephalopathy in *LAMB1*, including a homozygous substitution of cysteine for arginine (p.Cys460Arg) at the same amino acid residue as the nonsense variant identified here (Yasuda et al., 2020).

*LAMC1* is another laminin subunit which has recently been associated with familial CSVD in a UK-based cohort (Cho et al., 2024). We identified three rare, likely deleterious variants in *LAMC1*, one of which was also identified in the UK-based cohort (p.Pro321Ser) (Cho et al., 2024). *LAMC1* is known to interact with the laminin subunits *LAMB1* and *LAMA1* to form laminin 111, an essential component of the vascular basement membrane (Halder et al., 2023). Laminin 111 is known to self-organise with collagen IV, of which COL4A1 and COL4A2 are causative of monogenic CSVD (Kuo et al., 2012). Further, ablation of astrocytic laminin has been shown to impair vascular smooth muscle cell development, indicating a strong mechanistic role for *LAMA1* and other laminin subunits as causal of CSVD (Chen et al., 2013).

Several other key matrisomal genes were identified as significant in this study. Variants in *COL7A1* encode a collagen subunit and cause several forms of epidermolysis bullosa, an ECM disorder resulting in subepidermal blistering, yet have also been associated with neurological disorders such as Chiari malformation (Shinkuma, 2015; Urbizu et al., 2021). *MMP9*, a matrix metalloproteinase (MMP) that regulates extracellular matrix remodelling has been implicated in blood–brain barrier disruption, ischemic stroke, and white matter injury (Dong et al., 2009; Turner & Sharp, 2016). MMPs may also play a role in the pathophysiology of Alzheimer’s disease, with increased plasma levels of MMP9 identified in Alzheimer’s disease patients (Lorenzl et al., 2003).

*HMCN1* is a fibulin family ECM protein involved in cell adhesion and has been associated with age-related macular degeneration (Fisher et al., 2007). *HMCN1* is highly expressed in vascular smooth muscle cells of the cerebral arteries, a key cell type dysregulated in monogenic CSVD (Yu et al., 2024). *TNC* is another ECM glycoprotein that is expressed during vascular remodelling (Imanaka-Yoshida et al., 2014). The upregulation of this protein in post-acute diffuse traumatic brain injury suggests it may be associated with a regenerative response in neural or vascular tissues in the brain (Clancy et al., 2014; Griffiths et al., 2020). Additionally, plasma protein levels of TNC have been associated with WMHs in CSVD (Caro et al., 2025). The identification of multiple significant matrisomal protein-encoding genes highlights the role of ECM dysregulation in CSVD pathogenesis and the need for further characterisation as to their mechanistic role in these diseases.

*TENM4*, encoding teneurin transmembrane protein 4, is involved in oligodendrocyte development and central nervous system myelination, and is known to cause hereditary essential tremor (Hor et al., 2015). Mutations in this gene have also been associated with an increased risk of Parkinson’s disease (Pu et al., 2020). The high expression in neural tissues and role in myelination provides a plausible mechanism for *TENM4* as causative of white matter abnormalities and gait disturbance which is often seen in monogenic CSVD.

*TTN* encodes the sarcomeric protein titin which is a critical structural component of striated muscle and is frequently mutated due to its large size (Granzier & Labeit, 2025). *TTN* is known to cause a spectrum of myopathies including dilated and hypertrophic cardiomyopathies and is primarily expressed in skeletal and cardiac muscle (Loescher et al., 2021). Titin plays a role in regulating arterial stiffness through differential expression of titin isoforms in VSMCs, potentially implicating a role in CVSD through dysregulation of vascular compliance (Zhu et al., 2025).

In this study, we used a large cohort of clinically suspected CSVD patients, alongside a large control cohort free of neurological disease or dysfunction at an advanced age, to detect disease associations. However, segregation analysis of candidate variants was unable to be performed due to an absence of consented family members. Whole genome sequencing may also improve our ability to detect variants outside the coding regions which may play a role in CSVD. Extension of our findings within independent datasets may also be a valuable tool to more clearly define the role of implicated gene variants from our study. Finally, to further investigate the role of these genes in the pathogenesis of CSVD, functional studies using cell lines or animal models may also be beneficial.

## Conclusion

This study identified 18 rare, likely disease-causing variants in eight genes implicated in CSVD, including a significant burden of both rare and predicted deleterious variants in *ABCC6*. Pathogenic variants were also detected in four patients for neurological conditions with overlap to monogenic CSVD, including frontotemporal dementia with amyotrophic lateral sclerosis and familial intracranial aneurysm. Two genes previously implicated in conditions with phenotypic overlap to CSVD (*NOTCH1, MYH11*) were also significantly associated with CSVD in this cohort. Finally, rare, likely deleterious variants in *TTN, TENM4*, *TNC, COL7A1, MMP9, HMCN1,* and *LAMA1* were found to be significantly enriched in the case cohort, suggesting a potential contributory or causal role in CSVD. Notably, several of these genes are involved in the matrisome, further implicating its role in the molecular pathogenesis of CSVD.

## Ethics Statement

The study was approved by the Human Research Ethics Committee of the Queensland University of Technology (approval number: 14000007416) with appropriate written consent obtained from patients and in accordance with the Declaration of Helsinki. Diagnostic genetic testing was conducted under National Association of Testing Authorities (NATA) accreditation (accreditation number 14979).

## Conflicts of Interest

The authors have no relevant financial or non-financial interests to declare that are relevant to the contents of this study.

## Data Availability

All data produced in the present study are available upon reasonable request to the authors.

## Acknowledgements

The authors would like to express their thanks to each patient for providing consent to participate in this work.

## Funding

The authors disclose receipt of the following financial support for this article’s research, authorship and publication. This work was supported by the Australian National Health and Medical Research Council [NHMRC-APP1122387] (LRG), by infrastructure purchased with Australian Government EIF Super Science Funds as part of the Therapeutic Innovation Australia—Queensland Node project (LRG); and by a Queensland University of Technology Postgraduate Research Award [QUTPRA].

## Supplemental Material

Tables S1-9

Figure S1

## Notes

### Competing Interest Statement

The authors have declared no competing interest.

