## Supplementary figures and images for "Whole Exome Sequencing of Suspected Monogenic Cerebral Small Vessel Disease Patients reveals Novel Gene Associations"

### Supplementary Figure 1

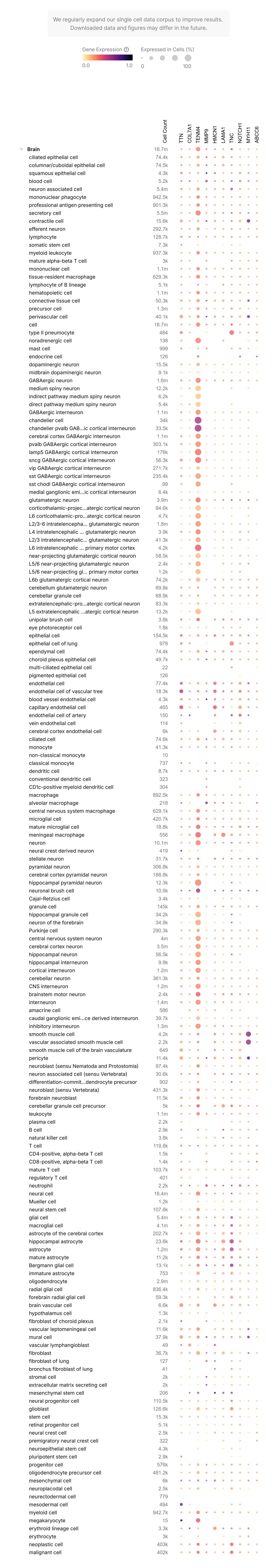
